# Representing Physician Suicide Claims as Nanopublications

**DOI:** 10.1101/2020.05.16.20101881

**Authors:** Tiffany I. Leung, Tobias Kuhn, Michel Dumontier

**Affiliations:** Faculty of Health, Medicine, and Life Sciences, Maastricht University, Maastricht, The Netherlands; Department of Computer Science, VU University Amsterdam, Amsterdam, The Netherlands; Institute of Data Science, Maastricht University, Maastricht, The Netherlands

## Abstract

**Motivation:** In the poorly studied field of physician suicide, various factors can contribute to misinformation or information distortion, which in turn can influence evidence-based policies and prevention of suicide in this unique population. Here, we report on the use of nanopublications as a scientific publishing approach to establish a citation network of claims drawn from a variety of media concerning the rate of suicide of US physicians. Our work integrates these various claims and enables the verification of non-authoritative assertions, thereby better equipping researchers and to advance evidence-based knowledge and make informed statements in the advocacy of physician suicide prevention.

## 1 INTRODUCTION

Nanopublications are “core scientific statements with associated context” (Groth et al., 2010). That is, scientific findings can be published as minimal pieces for computer interpretation, enabling nanopublications to cite other nanopublications unambiguously and reliably (Kuhn and Dumontier, 2014). Furthermore, they are self-contained in that they contain scientific assertions and their provenance information and metadata; nanopublications can then be given trusty URIs for verification of the digital artifact and also its entire reference tree (Kuhn and Dumontier, 2014). The infrastructure allows creation of citation, claim, and argumentation networks in which scientific statements are identified, connected, and verified (Groth et al., 2010; Clark et al., 2014; Clark, 2015).

In application, the use of nanopublications to represent scientific assertions in biomedical literature is not new. For example, the genetic basis for disease pathophysiology from Dis-GeNET has been mapped as nanopublications (Queralt-Rosinach et al., 2014). An Alzheimer disease research network built a web research community that organized research findings in an annotated knowledge base (Clark and Kinoshita, 2007). Applications largely involve datasets from the life science domains, including data on diseases, genes, proteins, drugs, biological pathways (Kuhn et al., 2018).

In the field of physician suicide, disparate research, opinion, and position statements have been published in scholarly literature, with more than 60% of such literature published in the last twenty years alone (Leung et al., 2019). Physician suicide has been reported in at least 37 countries and many risk factors for suicidal behavior that affect the general population, such as inadequately diagnosed or treated mental health disorders or substance use disorders, also apply to physicians. More controversially, various unique risk factors have been suggested, including specialized knowledge of human physiology, easier access to lethal means of self-harm, personality traits selected for in the physician training pathway, specialty of practice, and legal or licensing issues unique to the medical field (Leung et al., 2019).

Physician suicide is a serious issue for the medical workforce globally and maximally leveraging available evidence towards prevention. Yet, even foundational information about the incidence of physician suicide remains poorly understood. In previous work, a claim network was manually constructed to trace the provenance of an often-cited claim that 300 to 400 U.S. physicians die by suicide annually, which suggested that claim distortion and propagation of such misinformation about physician suicide incidence occurs in published literature (Leung et al., 2020). Nevertheless, this popular claim persists, most often stated as a variant of “300 to 400 U.S. physicians die by suicide annually.” A similar approach to identify and trace citation distortion had previously been done regarding a specific scientific claim about Alzheimer’s disease (Greenberg, 2009).

As literature about physician suicide is growing in parallel with the growth of scientific literature overall, this offers a unique opportunity to begin building core infrastructure to facilitate community learning, in a verifiable manner, about physician suicide. Such learning, founded on verifiability and reliability of available data, could support the needed vigilance of researchers, advocates, policymakers, and medical community in overcoming misinformation and information distortion about physician suicide.

In this paper, we aim to create nanopublications from assertions relating to physician suicide incidence. This is a proof-of-concept for applying semantic web infrastructure to physician suicide research. To our knowledge, no such application to this field has previously been done. Facilitating the integration, interoperability, and findability of high-quality research on physician suicide would benefit evidence-based policies and interventions in suicide prevention among physicians.

## 2 METHODS

### 2.1 Data sources

A previous scoping review of the literature about physician suicide identified articles that commented on or investigated suicidal behaviors of physician populations, including students, postgraduate trainees, and practicing physicians (Leung et al., 2019). A subset of articles from the literature search were identified that made an assertion (claim) about the annual rate of U.S. physicians who die of suicide. Additional articles published between August 2019 and March 2020 have been identified and manually added to the article set used for this study.

Manual data extraction was performed by one author (TIL) to collect article (or resource) type, title, authors, DOI or HTTP URI, publication year, claim (about annual physician suicide rate), data of last access of the article (e.g. for a webpage), and citations supporting the claim. Data was extracted into a spreadsheet that was then used to create nanopublications. For websites, a version of the website with a date nearby last access date was retrieved for data extraction.^1^ If a claim was available, then this text was extracted as the claim; if none, then the nanopublication included a comment, “No apparent claim of annual physician suicide rate”; if no archived version of the website was available, then the nanopublication included a comment, “Unverified claim of annual physician suicide rate present.” A nanopublication was created for each different cited version of website.

### 2.2 Data Structure

Each nanopublication consists of three components: the assertion, provenance, and publication information.^2^ Following the nanopublication model of Groth et al, the steps taken to create a nanopublication for each claim about physician suicide incidence involved:

1. The assertion: Represented as a set of triples: subject is the local article/resource identifier, which is linked via creator, date, identifier, title, type, citation(s) and comment.
2. The provenance: Each assertion is linked to the creator (annotator), who is identifiable by an ORCID account.
3. The publication information: Each nanopublication contains: a timestamp, the creator, link to the template, and pubkey plus signature.

We created a literature-based claim template^3^ to specify these fields and values, and provide mappings to semantic types and relations using RDFS, Nanopublication ontology, the Fabio ontology for document types, the Provenance, Authoring and Versioning (PAV) ontology for provenance, and the Semanticscience Integration Ontology (SIO) for citations.

### 2.3 Creating nanopublications

A nanopublication was created for each article or resource using Nanobench with the literature-based claim nanopublication template (illustrated in Figure 1). Nanobench is a Java based end-user tool that allows for browsing and publishing of nanopublications. By connecting to the decentralized nanopublication network (Kuhn et al., 2016), users can see other people’s nanopublications and publish their own via forms generated from specific templates, which are themselves defined and published as nanopublications. All published nanopublications are digitally signed and linked to the user’s ORCID account.^4^ A nanopublication index was then created containing all created nanopublications.

**Figure 1.**
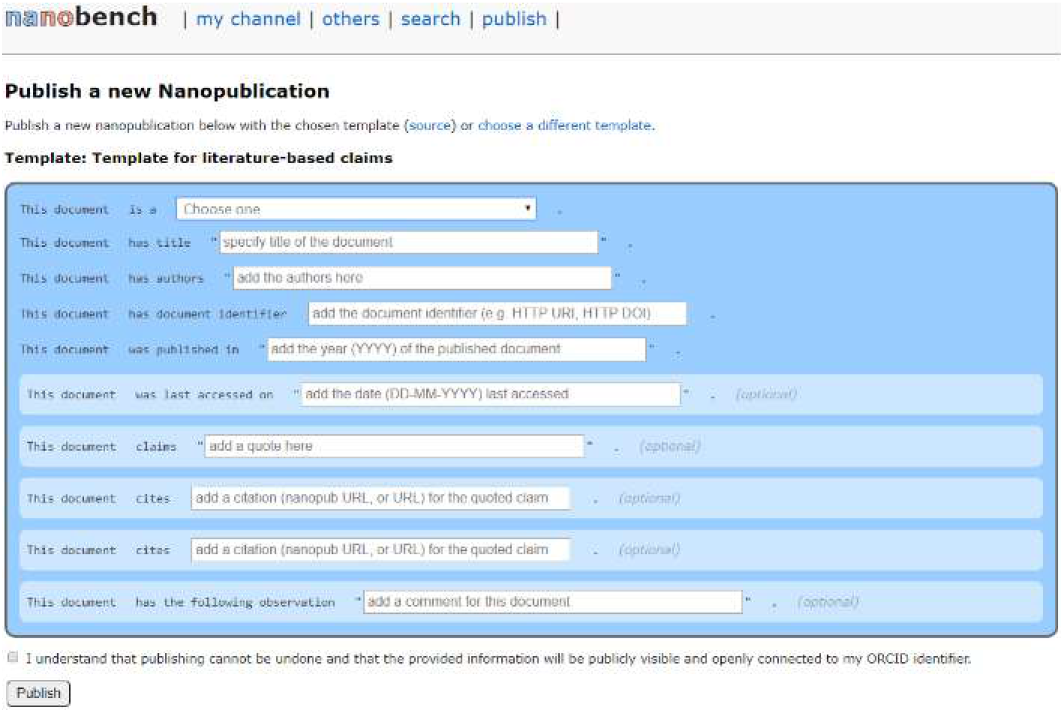
Nanobench template for literature-based claims.

## 3 RESULTS

A set of 49 claims concerning the rate of US physician suicide was represented as nanopublications. These claims were first manually curated into a spreadsheet^5^, subsequently published as individual nanopublications, and aggregated into a nanopublication index^6^. Figure 2 shows a chain of nanopublished claims.

**Figure 2.**
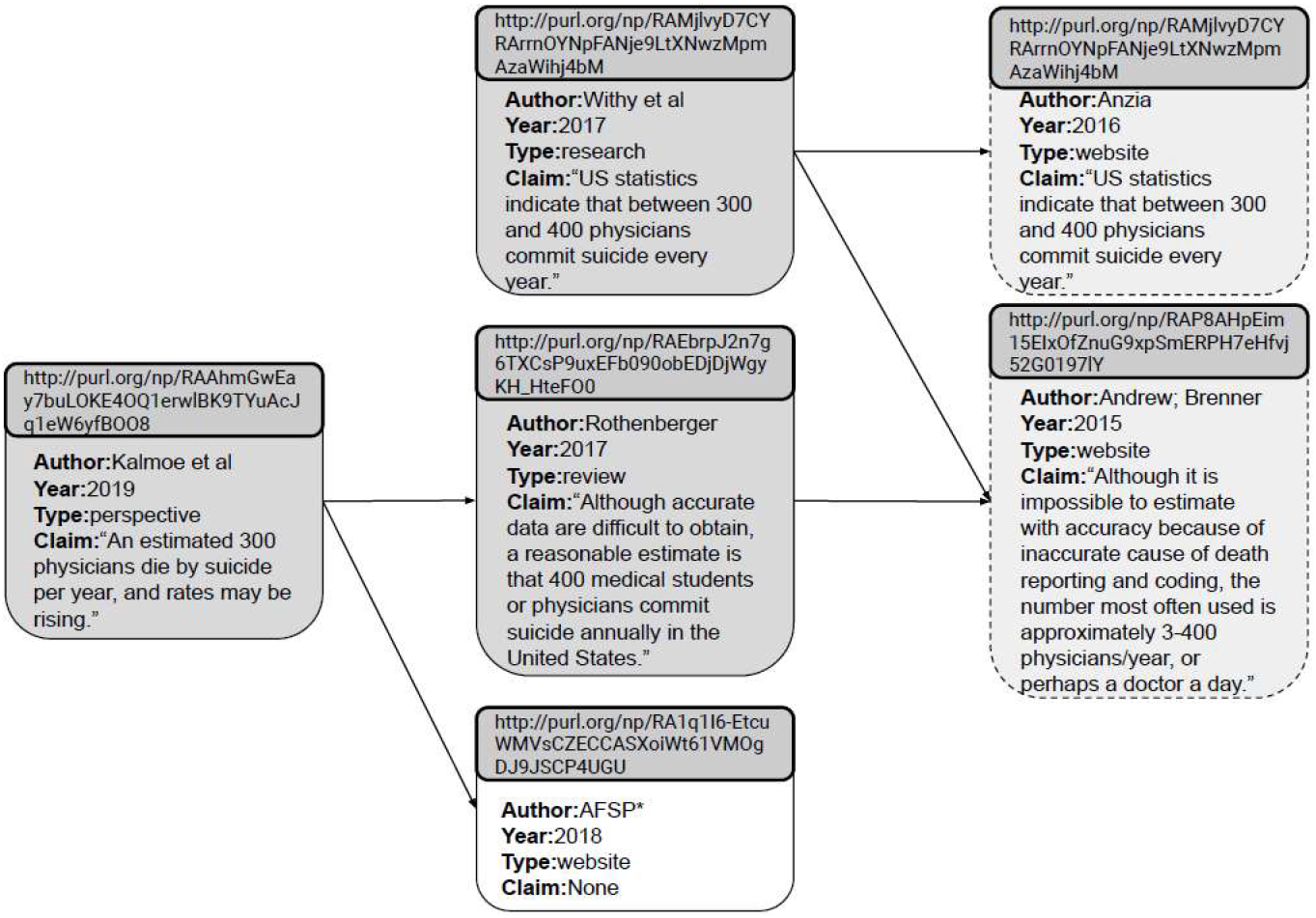
Nanopublications linked by their claims, if made, and nanopublications cited as source of the claim, if available. Nanopublications appearing in light gray with dashed represents an article or resource that states a claim about annual physician suicide rate but provides no further references. *AFSP = American Foundation for Suicide Prevention.

Analysis of the claim network (Table 1) revealed that (i) the network is not fully connected, (ii) no single primary source of the claim could be identified, and (iii) all end-point citations either had a claim with no further citation, no apparent claim, or could not be accessed to verify the claim.

The nanopublication strategy enabled the capture of variant claims published on a website. The website for the American Foundation for Suicide Prevention (AFSP) was cited 6 times between 2011 and 2018. Surprisingly, while only the 2018 version of the website could be retrieved, it contained no apparent claim of annual physician suicide rate. In addition, we found examples in which the claim itself changed over time. A 2015 version of the website published by Medscape stated, “*It has been reliably estimated that on average the United States loses as many as 400 physicians to suicide each year (the equivalent of at least one entire medical school)*.” However, a 2018 version of the website stated differently, “*Although it is impossible to estimate with accuracy because of inaccurate cause of death reporting and coding, the number most often used is approximately 3–400 physicians/year, or perhaps a doctor a day.*”

## DISCUSSION

This study has important implications. The findings of this study emphasize the importance of integrating scientific literature, especially individual scientific claims, in a reliable and verifiable manner. Creating nanopublications to represent articles’ claims that “300 to 400 U.S. physicians die by suicide annually” demonstrates that this is a poorly supported yet frequently stated claim (Table 1), in line with previous findings (Leung et al., 2020). However, this study builds on previous work by applying nanopublication infrastructure to the articles and claims they make. Nanopublications allow for continued claim tracing and verification, including, for example, accounting for versioning. Interestingly, different website versions may even differ in their assertions of the claim, as seen in the Medscape article. Nevertheless, the nanopublication approach remains to be adopted in broader scientific publishing in medicine, and especially in publishing about physician mental health.

**Table 1.**
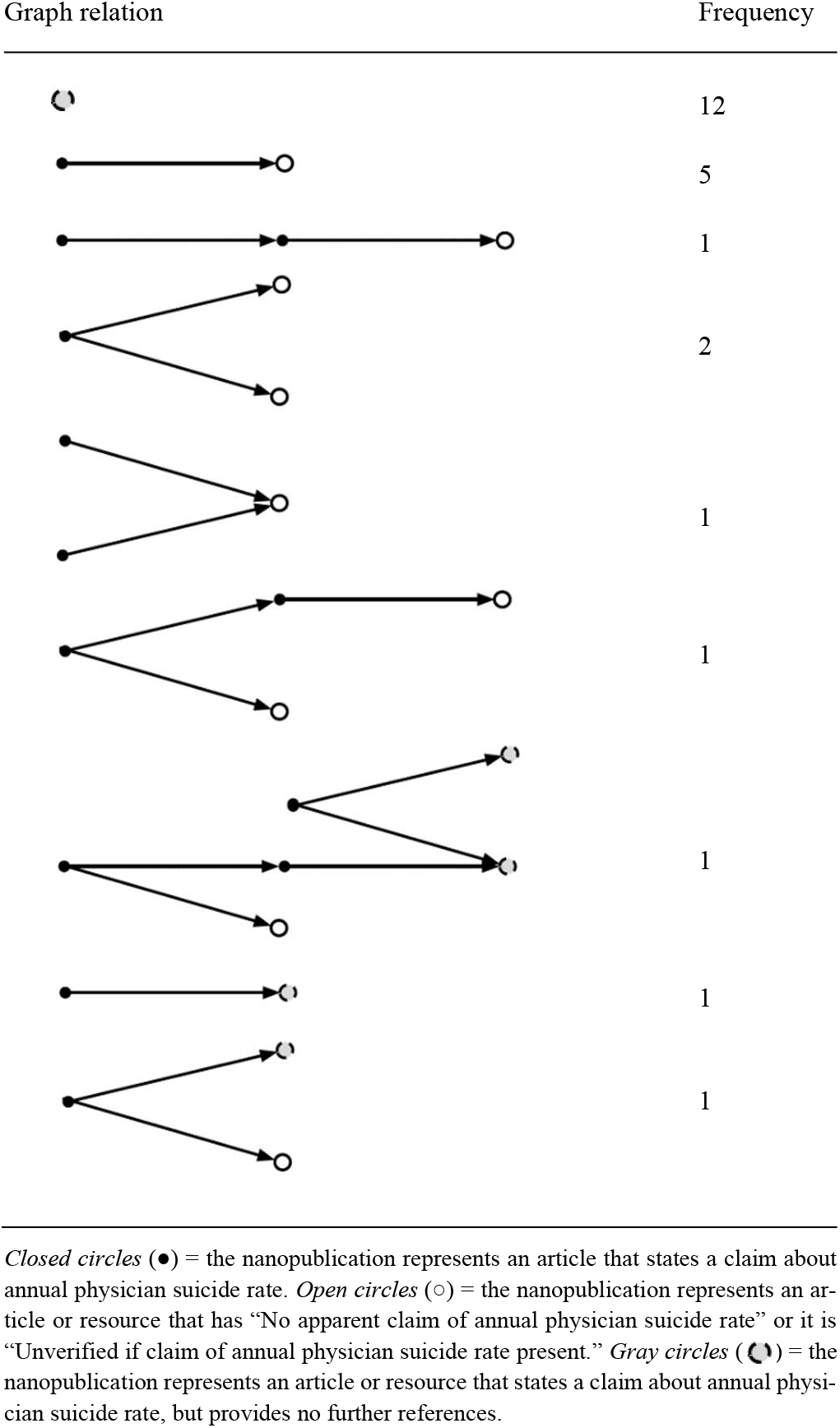
Extracted sub-graphs from the physician suicide claim network. Graph relation Frequency.

One limitation is that nanopublication quality depends on a minimum set of community agreed-upon annotations (Groth et al., 2010). In the study of physician suicide, no such community standards exist; standardized data collection about suicidal behaviors or suicide deaths is limited (Leung et al., 2019). However, this could also be an opportunity to develop such standards, driving the application of nanopublication in this field from the ground up.

Another limitation is that the claim network contains only verbatim claims about the annual physician suicide rate. The first study estimating incidence from two years of obituary data from a medical professional organization was published in 1968, reporting a crude annual suicide rate of 38.4 per 100,000 physicians (Craig and Pitts, 1968). Since then systematic reviews or meta-analyses have sought to aggregate data from other observational studies estimating incidence (Schernhammer and Colditz, 2004; Blacker et al., 2019; Dutheil et al., 2019; Duarte et al., 2020). Most studies about suicide incidence should report a suicide mortality rate (SMR), which is the number of deaths by suicide per 100,000 person-years, and physician SMRs have yet to be nanopublished. Further work is needed to represent all available data on physician suicide, beyond focusing on the single claim studied here. Representing additional data as nanopublications, including incidence data, risk factors, demographics, and other contextual information, may offer an even richer graph of existing knowledge about physician suicide to enable more rapid learning about the field.

## Data Availability

Data curated and used for this study are available online.

https://doi.org/10.6084/m9.figshare.12221870

## Footnotes

1 https://archive.org/web/

2 http://nanopub.org/

3 http://purl.org/np/RAqWlNPJt3Eb4HkmPCpjaiRHGCzKIZag6cBNMkG8nxu6I

4 https://github.com/peta-pico/nanobench/

5 https://doi.org/10.6084/m9.figshare.12221870

6 http://purl.org/np/RAzPytdERsBd378zHGvwgRbat1MCiS7QrxNrPxe9yDu6E

